# Health outcomes after national acute sleep deprivation events among the American public

**DOI:** 10.1101/2024.10.29.24316369

**Authors:** Neil J. Kelly, Rahul Chaudhary, Wadih El Khoury, Nishita Kalepalli, Jesse Wang, Priya Patel, Irene N. Chan, Haris Rahman, Aisha Saiyed, Anisha N. Shah, Colleen A. McClung, Satoshi Okawa, Seyed Mehdi Nouraie, Stephen Y. Chan

## Abstract

**Background:** Sleep is increasingly recognized as essential to human health, yet the adverse health consequences of acute sleep deprivation are unknown. Using actigraphic, genomic, and health data from the *All of Us* (*AoU*) Research Program, we characterized the detrimental health consequences of acute sleep deprivation in the American public.

**Methods:** LOESS smoothing was performed on sleep estimates from Fitbit users (N = 14,681) between June 1, 2016 and July 1, 2022. Dates when population minutes slept were less than the 90% confidence interval of the LOESS regression were named acute sleep deprivation events (ASDEs). Phenome-wide disease incidence among the *AoU* population (N = 287,012) in the 10 days post-ASDE was compared to a preceding reference period by McNemar test. Circadian rhythm and sleep duration-associated SNPs were screened to identify genotypes associated with shorter ASDE sleep duration. Influences of sleep and circadian genotype on post-ASDE influenza risk were modeled using binomial family generalized estimating equations.

**Findings:** We identified 32 ASDEs spanning political and non-political events. A phenome-wide screen found increased risk of influenza (OR = 1.54 [1.40, 1.70], *P*-value = 1.00 x 10^-18^) following ASDEs. 56 SNPs were associated with decreased sleep duration on ASDEs. Higher quantiles of ASDE-related SNP genotype burden were associated with less ASDE sleep duration and a greater risk of influenza-associated healthcare visits.

**Interpretation:** National political and non-political events are associated with acute sleep deprivation and greater influenza risk which is amplified by sleep genotypes. These findings should inform public health vigilance surrounding major national events.

## Introduction

Sleep is essential and foundational for human health^1^. Yet insufficient sleep duration is a common problem affecting nearly one third of all U.S. adults which some studies suggest is becoming more pronounced over time^2,3^. The driving causes behind sleep deprivation are numerous, spanning individual, career, social, and cultural factors^1,4^.

Chronic sleep deprivation has been linked to myriad negative health consequences including neurocognitive impairment^5^, mental health and behavioral disorders^6^, altered metabolism^7^, immune dysregulation^8,9^, cardiovascular disease^10,11^, and all-cause mortality^12^. Contrasted with chronic sleep deprivation, our understanding of the health impacts of acute sleep deprivation is limited. Based on small studies in laboratory settings, acute sleep deprivation is thought to have adverse effects on neurobehavioral, endocrine, and immune physiology^13^. Recently, a small study detected significant changes in the serum proteome after just a single night of sleep deprivation^14^. However, the impacts of these transient changes on human disease are largely unknown. In part, this knowledge deficit may be explained by the difficulty of acute sleep deprivation studies in humans, which traditionally involve laboratory-based studies of healthy volunteers in controlled conditions. In other cases, studies are limited to small populations where sleep is monitored through diaries or wearable biosensors^15^.

The *All of Us* (*AoU*) Research Program released Fitbit-based daily sleep estimates for a subset of its >413,000 participants. This resource has revealed phenomic associations between chronic sleep patterns and disease^16^. Here, we sought to leverage *AoU* to identify health events associated with acute sleep deprivation. First, by Fitbit-based sleep monitoring, we identified dates that coincided with major political and non-political events and displayed acute sleep deprivation largely shared across the *AoU* subpopulation. Such a strategy bypassed any study confounders related to the heterogeneity of sleep deprivation patterns observed at the individual level across the calendar year. Second, in an unbiased phenome-wide analysis, we identified an increase in influenza incidence following population-level acute sleep deprivation. Finally, we incorporated *a priori* knowledge of sleep genomics to decouple the effects of sleep and circadian disturbances from other related stressors. By this approach, we found that acute sleep deprivation increases the risk of influenza, and that this risk is amplified by genotypes associated with shorter sleep duration.

## Methods

### Data availability

*AoU* data is available to authorized users of the *All of Us* Research Program’s Controlled Tier Dataset 7 on the Researcher Workbench at https://www.researchallofus.org/.

### All of Us

This study used data from the *All of Us* Research Program’s Controlled Tier Dataset 7. The study period encompassed June 1, 2016 through July 1, 2022. No participants were excluded from the study.

### Television viewership

Television viewership data was obtained from archived online reports of Nielsen cable and national broadcast ratings^17^.

### Google Trends

Google Trends is an online tool which records archival internet search volume^18^. The top 3 Google Trends search queries in the United States were annotated for the day prior to each ASDE.

### Time zones

5-digit U.S. ZIP code coordinates were obtained from the U.S. Census Bureau ZIP Code Tabulation Areas Gazetteer File^19^ of 2020 Census tabulation blocks. In order to identify coordinates for the 3-digit ZIP codes supplied by the *AoU* Controlled Tier Dataset, the constituent 5-digit ZIP code coordinates (latitude and longitude) were averaged. Time zones of the resultant coordinates were obtained from the Python implementation of *googlemaps*.

### State political partisanship

The Cook Partisan Voter Index (Cook PVI^SM^) measures a state’s lean towards the Republican (R) or Democratic (D) Party in U.S. presidential elections (e.g. R+1 indicates a one percentage point Republican lean). 2022 Cook PVI^SM^ was obtained from publicly available data^20^. For the generation of quartiles, leans were expressed as negative or positive numbers depending on party.

### Time-weighted average positive influenza tests

National weekly positive influenza tests (Influenza A and Influenza B) from clinical laboratories were obtained from the U.S. Centers for Disease Control and Prevention’s *FluView* tool for the 2016-2017 through 2021-2022 seasons^21^. Positive tests were averaged over all seasons by week of the year. The average positive tests corresponding to a time period were determined by averaging the weekly positive tests for the week of the year associated with each day of the period and multiplying by the number of weeks in the period.

### Acute Sleep Deprivation Event (ASDE) determination

Daily weekday minutes slept were averaged for all *AoU* participants with available sleep data. Locally estimated scatterplot smoothing (LOESS) regression of minutes slept, as well as confidence intervals, were generated using the Python package *tsmoothie* v1.0.5 with a smoothing fraction of 0.1 and one iteration. Because of our one-sided focus on sleep deprivation, dates with average minutes slept less than the 90% confidence interval of the LOESS regression were considered ASDEs.

### Post-ASDE and reference period selection

Post-ASDE periods were defined as the ten days beginning on the ASDE. If a post-ASDE period overlapped with a subsequent post-ASDE period, they were pooled into a combined period beginning on the earliest of the overlapping ASDEs and ending on the tenth day of the latest ASDE period. Reference periods were chosen for each pooled post-ASDE period as the latest period of the same duration beginning on the same weekday as its corresponding post-ASDE period which did not overlap with a post-ASDE period. For the separate comparison of political post-ASDEs, reference periods were selected as the period of the same duration beginning on the same weekday as its corresponding post-ASDE period which was closest to 365 days after the corresponding ASDE.

### AoU diagnoses

Data were organized and annotated using the Observed Medical Outcomes Partnership (OMOP) common data model. Conditions occurring within post-ASDE or reference periods were considered. A condition was considered to have occurred if either the condition or one of its 1^st^ degree descendants occurred in a period. For the phenome-wide analysis, conditions which had been previously associated with the participant prior to the period start date were excluded so as only to examine new diagnoses. In order to focus our search on clinical disorders rather than symptoms, we limited our analysis to conditions with an ancestor of (*1*) “Mental Disorder” (ID 432586) or (*2*) “Disorder of Body System” (ID 4180628) and either “Disorder of nervous system” (ID 376337), “Disorder of endocrine system” (ID 31821), “Disorder of digestive system” (ID 4201745), “Disorder of cardiovascular system” (ID 134057), “Disorder of musculoskeletal system (ID 4244662), “Disorder of the genitourinary system (ID 4171379), “Disorder of respiratory system” (ID 320136), “Disorder of auditory system” (ID 4176644), “Disorder of lymphatic system” (ID 440363), “Red blood cell disorder” (ID 432739), “Hereditary disorder by system” (ID 4180158), or “Visual system disorder” (ID 4134440). For analyses of influenza specifically, including analysis of political events, Thanksgiving, and generalized estimating equations, we allowed for the possibility that an individual may contract influenza in multiple years; therefore, the diagnosis of influenza was considered to have occurred in a given period if it was diagnosed in that period and had not been diagnosed in the preceding 180-day period.

### Single nucleotide polymorphisms (SNPs)

SNPs associated with circadian rhythm (N = 133 total, N = 126 in *AoU* genomics records) and sleep duration (N = 960 total, N = 895 in *AoU* genomics records) were obtained from the GWAS Catolog^22^ using the trait identifiers GO_0007623 and EFO_0005271, respectively. Genotypes, sex ploidy, and genomic ancestry were obtained from the *AoU* workbench^23^. Minor allele frequencies (MAFs) were calculated for each SNP allele based on their frequencies in the *AoU* genomics population (N = 243,480). Alleles with MAF > 5% and allele combinations present at least 20 participants in the *AoU* genotyped Fitbit population (N = 1,926 allele combinations) were included.

### Software

Summary statistics were plotted in GraphPad Prism v10.2.0 for Windows (GraphPad Software, Boston, MA). Statistical tests were performed in Python v3.10.12 or Graphpad Prism as specified. *P*-value < 0.05 was considered statistically significant unless otherwise noted.

### Sleep deprivation comparison

Daily minutes slept were smoothed using LOESS regression with a smoothing fraction of 0.1 and one iteration for each subgroup (e.g. male and female, political lean quartile). Sleep deficit was calculated as the difference between the subgroup’s mean sleep duration and its LOESS fit for each ASDE. Comparisons between subgroups were performed using by Mann-Whitney test (two subgroups) or Kruskal-Wallis test with Dunn’s multiple comparisons (three or more subgroups) using GraphPad Prism.

### Phenome-wide risk ratio analysis

Post-ASDE and reference periods were pooled to generate a single combined post-ASDE or reference period such that overlapping timeframes were not double counted. Diagnoses corresponding to fewer than 20 participants in either the combined (corresponding to all ASDEs) reference periods or ASDE periods were excluded per *AoU* data use policy. Comparisons of condition instances between time periods were conducted using the *mcnemar* implementation of McNemar’s test in the *scipy* package (v1.11.2)^24^. Odds ratio (OR) was estimated as the number of post-ASDE divided by the number of reference instances. 95% confidence intervals were calculated using the *scipy* binomial test *binom_test*. Bonferroni-adjusted *P*-values < 0.05 were considered statistically significant for the phenome-wide analysis.

### SNP sleep duration comparison

Average ASDE sleep duration was compared to that of the *AoU* population using the Mann-Whitney test (*mannwhitneyu* function of the Python package *scipy*). 95% confidence intervals were calculated using the *scipy bootstrap* function with default parameters. Given the *a priori* association of these SNPs with circadian rhythm and sleep duration^22^, multiple test correction was performed by the Benjamini-Hochberg procedure with a false discovery control level of α = 0.05 using the *scipy* method *false_discovery_control*. Homozygous SNP genotypes with a sleep ratio (vs. *AoU* population) less than 1.0 and adjusted *P*-value < 0.05 were considered short sleep genotypes. Genotyped *AoU* participants were assigned to quantiles based on their number of short sleep genotypes using the *pandas qcut* function in Python with 2 groups centered on the median. Heterozygous genotypes containing a significant allele were counted as 0.5 short sleep genotypes. ASDE sleep duration comparison between quantiles was performed by Mann-Whitney test in Graphpad Prism.

### Generalized estimating equations (GEEs)

GEEs of the binomial family distribution with logit link function, exchangeable dependence structure, and robust standard errors were used to calculate relationships between hospitalizations, time period (pre- or post-ASDE), short sleep genotype quantile, sex chromosome ploidy, genomic ancestry, average positive influenza tests, and age at ASDE. Covariates were selected based on prior knowledge^25-27^. GEE analysis was performed using *statsmodels* v0.14.2^28^.

### Role of the Funding Source

The study sponsors had no role in study design; in the collection, analysis, and interpretation of data; in the writing of the report; and in the decision to submit the paper for publication.

## Results

### Acute sleep deprivation events in the AoU population

Using Fitbit-based estimates of average daily weekday minutes slept in the cohort population (N = 14,681 Fitbit users, **Table 1**), we modeled expected daily minutes slept based on LOESS regression (**Figure 1A**). Next, to identify dates where average minutes slept were significantly less than expected, we identified 32 dates where the average minutes slept was less than the lower bound of the 90% confidence interval of the LOESS regression. We defined these dates as acute sleep deprivation events (ASDEs) and annotated each date based upon temporal data from Google Trends^18^, television viewership^17^, and curated current events^29^ (**Table 2, TableS1**). The ASDEs corresponded to major national political occasions, holidays, media events, and athletic competitions. On these dates, we found no significant differences in sleep deprivation based upon sex, age, state of residence political leaning, race, time zone, and education level, suggesting that acute sleep deprivation spans multiple segments of the population (**Figure 1, B-G**).

**Table 1.** Baseline characteristics of the AoU participants. Data are divided by all *AoU* participants (AoU) and those with whole genome sequencing (WGS). Within each category, data are divided into all participants (All), those with linked electronic health records (EHR), and those with linked Fitbit data after June 1, 2016 (Fitbit). Continuous variables are presented as median [interquartile range] and mean ± standard deviation. Categorical variables are presented as *n* (%).

|  | AoU |  |  | WGS |  |  |
| --- | --- | --- | --- | --- | --- | --- |
|  | All | EHR | Fitbit | All | EHR | Fitbit |
| N | 413,457 | 287,012 | 14,681 | 243,480 | 204,478 | 8,276 |
| Age (yrs) | 49 [33,61],<br>47.80±17.11 | 51 [35,62],<br>48.97±16.96 | 48 [34,60],<br>47.12±15.73 | 50 [34,62],<br>48.27±16.91 | 51 [35,62],<br>48.91±16.83 | 50 [35,61],<br>48.21±15.50 |
| **Sex** |  |  |  |  |  |  |
| Female | 249,565 (60.36) | 172,401 (60.07) | 10,056 (68.50) | 145,430 (59.73) | 123,857 (60.57) | 5,730 (69.24) |
| Male | 155,169 (37.53) | 108,739 (37.89) | 4,213 (28.70) | 93,019 (38.20) | 76,439 (37.38) | 2,317 (28.00) |
| Other/Unknown | 8,723 (2.11) | 5,872 (2.05) | 412 (2.81) | 5,031 (2.07) | 4,182 (2.05) | 229 (2.77) |
| **Race** |  |  |  |  |  |  |
| Asian | 14,215 (3.44) | 8,294 (2.89) | 460 (3.13) | 7,614 (3.13) | 5,751 (2.81) | 269 (3.25) |
| Black or African American | 78,504 (18.99) | 58,264 (20.30) | 731 (4.98) | 50,848 (20.88) | 40,745 (19.93) | 406 (4.91) |
| Other/Uknown | 91,580 (22.15) | 65,776 (22.92) | 1,554 (10.59) | 57,006 (23.41) | 46,257 (22.62) | 869 (10.50) |
| White | 229,158 (55.42) | 154,678 (53.89) | 11,936 (81.30) | 128,012 (52.58) | 111,725 (54.64) | 6,732 (81.34) |

**Table 2.** Acute sleep deprivation events. ASDE sleep deprivation was defined as the difference between the LOESS fit of average daily minutes slept and actual average minutes slept. Google Trends and television viewership are presented for the day prior to the ASDE.

| ASDE | Sleep Loss (minutes) | Notable U.S. Events | Top Google Trend | Most Viewed Television Program |
| --- | --- | --- | --- | --- |
| 07/29/2016 | 7.16 | Democratic National Convention | dunkin save | Americas Choice 2016 |
| 08/12/2016 | 6.17 | Summer Olympics | verge depre | Sum Olym Thu Prime |
| 09/27/2016 | 6.48 | Presidential Debate | arnold palmer | Big Bang Theory, The |
| 10/20/2016 | 8.31 | Presidential Debate | debate time | Presidential Debate |
| 11/03/2016 | 13.78 | World Series | world series game 7 | FOX World Series |
| 11/09/2016 | 43.64 | Presidential Election | election results 2016 usa | Election Night In America |
| 02/06/2017 | 6.25 | Super Bowl | snl sean spicer | FOX Super Bowl LI |
| 05/26/2017 | 6.04 | NBA & NHL Playoffs | gianforte | Big Bang Theory, The |
| 06/19/2017 | 6.13 | International Cricket Cup | happy fathers day 2017 | Celebrity Family Feud |
| 07/05/2017 | 6.72 | Independence Day | joey chestnut | Macy's 4th July Fireworks |
| 12/08/2017 | 6.87 | Thursday Night Football | lilac fire | NBC+NFLN Thu Nt Football |
| 12/25/2017 | 9.23 | Christmas Eve | norad | OT, The |
| 01/01/2018 | 16.43 | New Year's Eve | ufc 219 results | Primetime NYRE '18 |
| 02/05/2018 | 10.27 | Super Bowl | kylie jenner baby | Superbowl Game LII |
| 12/18/2018 | 6.16 | Monday Night Football | quizlet live | New Orleans/Carolina |
| 12/25/2018 | 10.93 | Christmas Eve | santa tracker norad | Denver/Oakland |
| 01/01/2019 | 14.83 | New Year's Eve | elizabeth warren | Primetime NYRE '19 |
| 05/20/2019 | 6.56 | Game of Thrones Finale | robert f smith | Game Of Thrones |
| 05/29/2019 | 6.84 | America's Got Talent Premiere | ohio tornado | America's Got Talent |
| 06/07/2019 | 6.55 | FIFA Women's World Cup | granger smith | Celebrity Family Feud |
| 12/20/2019 | 7.46 | Primary Debate | john dingell | Young Sheldon |
| 01/01/2020 | 11.62 | New Year's Eve | new year's eve | Primetime NYRE '20 |
| 03/12/2020 | 6.12 | COVID-19 Pandemic | josie harris | Masked Singer, The |
| 09/30/2020 | 9.06 | Presidential Debate | debate tonight | Presidential Debate |
| 10/23/2020 | 6.90 | Presidential Debate | what time is the debate tonight | Presidential Debate |
| 11/04/2020 | 23.87 | Presidential Election | election results #election2020 | FOX News Democracy 2020 |
| 11/06/2020 | 9.08 | Thursday Night Football | why is nevada taking so long | FOX+NFLN Thu Nt Football |
| 12/25/2020 | 11.19 | Christmas Eve | charles kushner | NBC Movie Special |
| 01/01/2021 | 11.12 | New Year's Eve | mf doom | Primetime NYRE '21 |
| 01/07/2021 | 8.79 | Election Certification | capitol building | BN/Mob Attacks US Capitol |
| 06/07/2021 | 6.37 | CONCACAF Soccer Final | tb joshua | 60 Minutes |
| 12/17/2021 | 6.53 | Thursday Night Football | todrick hall | 2021 WK15 KC VS. LAC |

**Figure 1.**
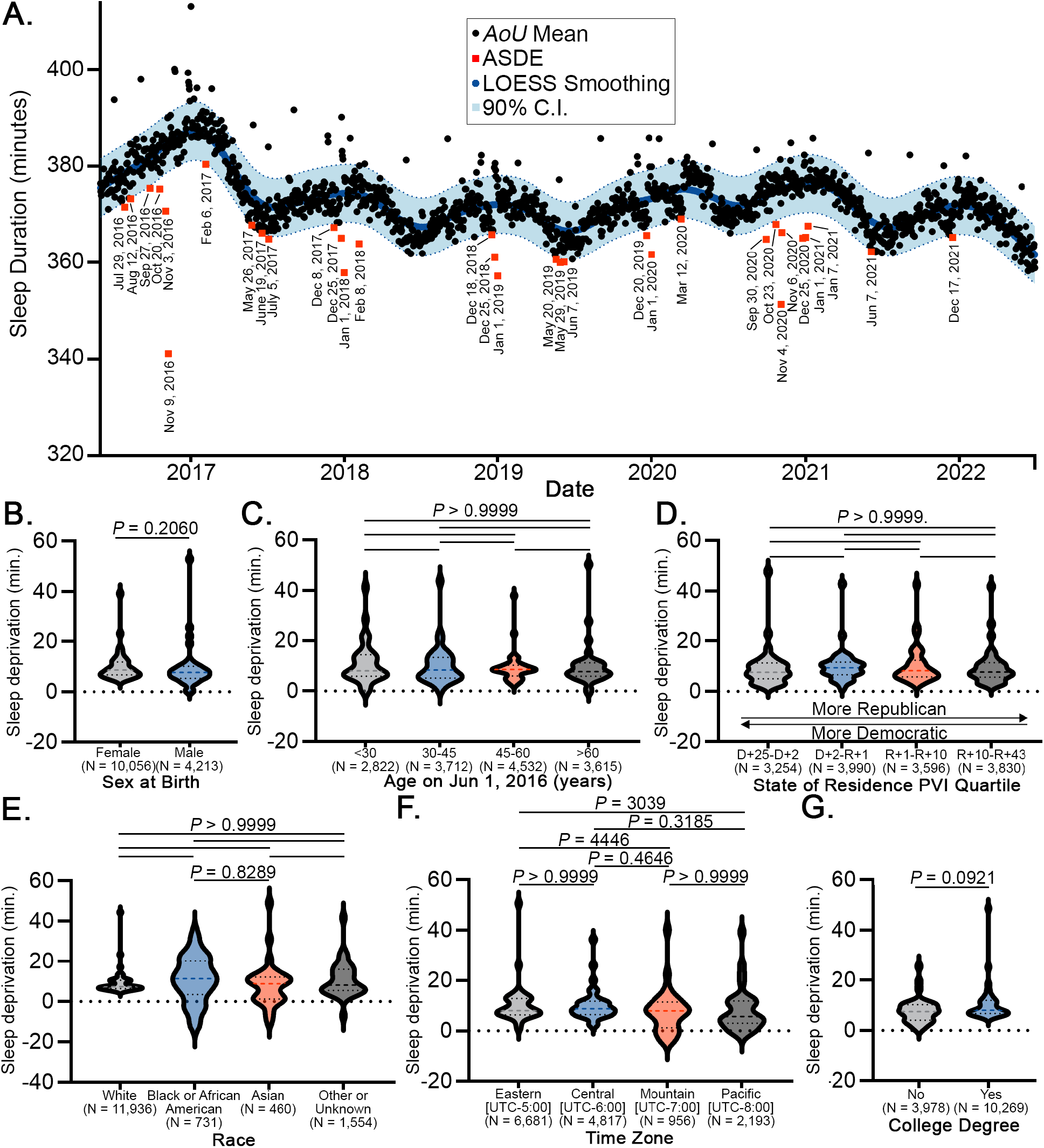
Acute sleep deprivation events across the AoU population (Jun 1, 2016 – Jul 1, 2022). (**A**) Daily average minutes slept were fit to a LOESS regression. Dates below the 90% confidence interval (C.I.) were named ASDEs (N = 14,681). For each category of (**B**) sex at birth, (**C**) age range, (**D**) state political lean, (**E**) race, (**F**) time zone, and (**G**) education, LOESS smoothing was performed and average minutes slept on ASDEs were subtracted from the LOESS fit to determine minutes sleep deprivation. Dashed line on violin plot is median; dotted lines are interquartile range. Statistical comparisons were made by Mann-Whitney test (2 groups) or Kruskal-Wallis test with Dunn’s multiple comparisons (3+ groups). D: Democratic; R: Republican; UTC: Coordinated Universal Time. *P* < 0.05 was considered statistically significant.

### ASDEs are associated with increased influenza healthcare visits

In order to identify health outcomes associated with ASDEs, we conducted an unbiased phenome-wide analysis of the entire *AoU* population with linked electronic health record (EHR) data (N = 287,012, **Table 1**). Incident new diagnoses from any healthcare visit in the 10-day period following an ASDE were compared with those occurring in a preceding 10-day reference period (**Figure 2**). Overlapping post-ASDE periods were pooled (to N = 22 ASDEs) such that the period concluded 10 days following the latest ASDE. Reference periods (**Table S2**) were selected as the nearest preceding non-overlapping timeframe beginning on the same weekday as the ASDE. New diagnoses occurring in the pooled post-ASDE periods versus reference periods were compared by McNemar’s test, which identified a significantly increased risk of influenza (1,052 vs. 683, OR = 1.54 [1.40, 1.70], *P*-value = 1.00 x 10^-18^) and other related respiratory conditions in the pooled post-ASDE period (**Table S3**). Based on this finding, we directed our attention to the association of ASDEs and influenza risk.

**Figure 2.**
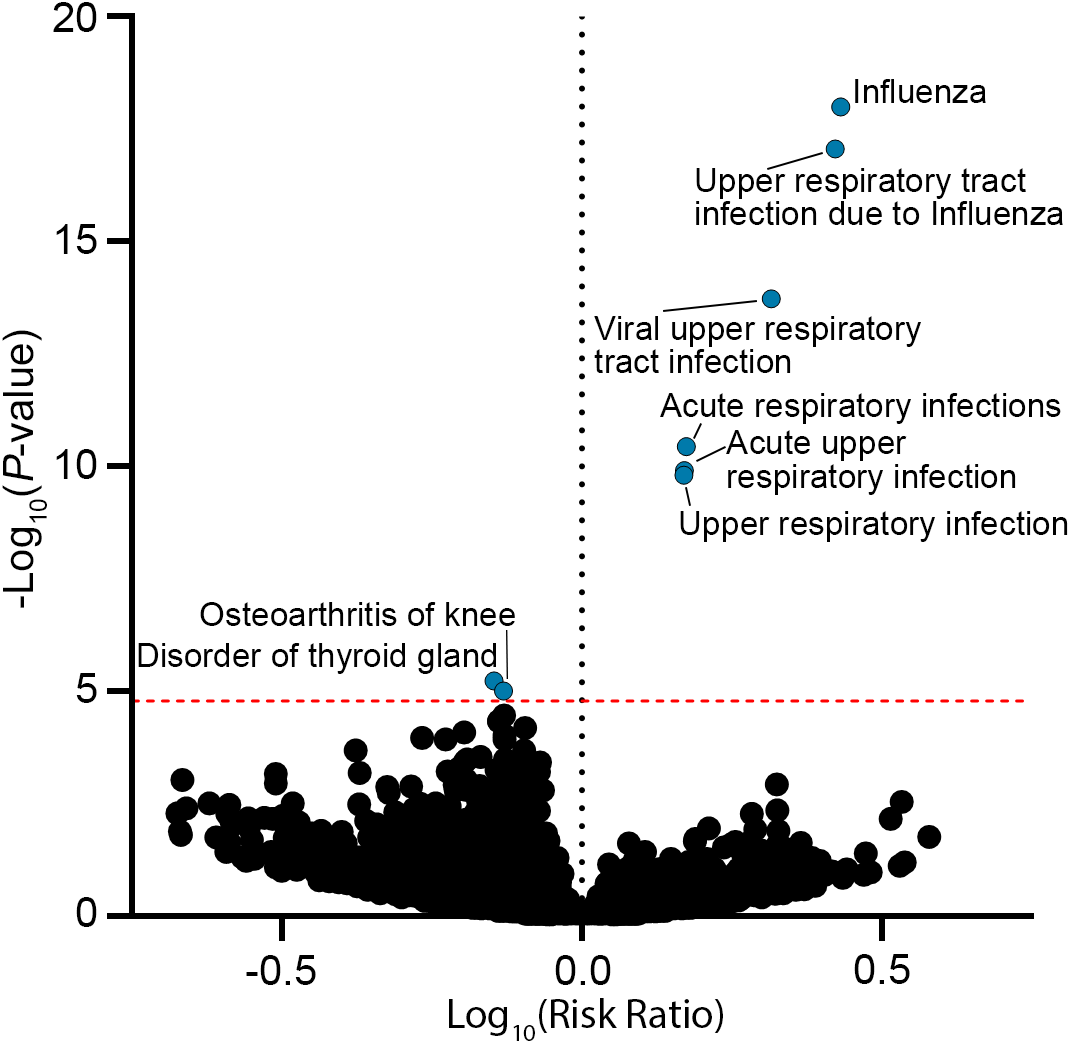
Influenza and acute respiratory infections are increased following ASDEs. Phenome-wide frequency of new diagnoses was compared between post-ASDE periods and reference periods by McNemar’s test in the *AoU* population (N = 287,012). Red dotted line represents Bonferroni-adjusted *P*-value threshold. Diagnoses with significant (adjusted *P*-value < 0.05) differences in risk ratio are annotated.

Because the duration of the ASDE effect is unknown, we only compared post-ASDE periods to preceding (as opposed to trailing) reference periods. However, to account for the strong seasonality of influenza incidence^30^, we leveraged the fact that national political events tend to occur in 4-year cycles. In this regard, we compared the incident influenza healthcare visits in the post-ASDE periods of national political events (see **Table 2**) to their incidence in a reference period exactly one year later (**Table S4**). Consistent with our hypothesis that ASDEs were associated with increased influenza risk, we observed an increased number of influenza-associated healthcare visits in the political post-ASDE periods as compared to the following year (166 vs. 111, OR = 1.50 [1.17, 1.92], *P*-value = 0.0012). Additionally, it is possible that confounding factors such as travel, stress, and anxiety associated with national events and holidays drive the observed signal. However, the post-Thanksgiving holiday period, which was not an ASDE but may have similar characteristics to other major U.S. holidays (**Table S4**), did not have a significant association with influenza healthcare visits compared to the preceding reference period (123 vs. 106, OR = 1.16 [0.89, 1.52], *P*-value = 0.2883). Taken together, these data suggest an association between ASDEs and influenza risk.

### Identification of short sleep genotypes

Given the apparent link between ASDEs and influenza risk, we hypothesized that individuals who are genetically predisposed to less sleep would be at greater risk of an influenza-associated healthcare visit after an ASDE. Misalignment of the circadian rhythm and the sleep homeostat, both of which are subject to genetic influence^31^, contribute to human disease manifestation across a broad range of organ systems. Therefore, we hypothesized that genetic predispositions in sleep duration modify the risk of post-ASDE incident disease. We first obtained lists of single nucleotide polymorphisms (SNPs) associated with either circadian rhythm (GO: 0007623) or sleep duration (EFO: 0005271) from the publicly-available genome-wide association study (GWAS) Catalog^22^. Using the genotyped segment of the *AoU* Fitbit cohort (N = 8,276), we identified common variants (minor allele frequency > 5%^32^, **Table S5**) for further study. For every homozygous common variant genotype present in at least 20 *AoU* Fitbit participants, we quantified the average sleep duration on the 32 ASDEs as compared to the entire *AoU* Fitbit population. Through this approach, we identified 56 SNP genotypes associated with less ASDE sleep than the *AoU* population mean (**Figure 3A, Table S6**), which we name “short sleep genotypes.” Participants were then binned into two quantiles according to their having at most (1^st^ quantile, N = 127,652) or greater than (2^nd^ quantile, N = 115,828) the median number, 28, of short sleep genotypes. Heterozygous genotypes containing a significant allele were counted as 0.5 short sleep genotypes. We observed a significant decrease in ASDE sleep duration between short sleep genotype quantiles (**Figure 3B**); of note, bin proportions differed between the overall population and the Fitbit population because the median of the overall population was used in binning. Interestingly, there was also a shift in baseline sleep duration between genotype quantiles (**Figure 3C**), indicating that ASDE-associated sleep deficits take place in addition to genetically predisposed sleep durations.

**Figure 3.**
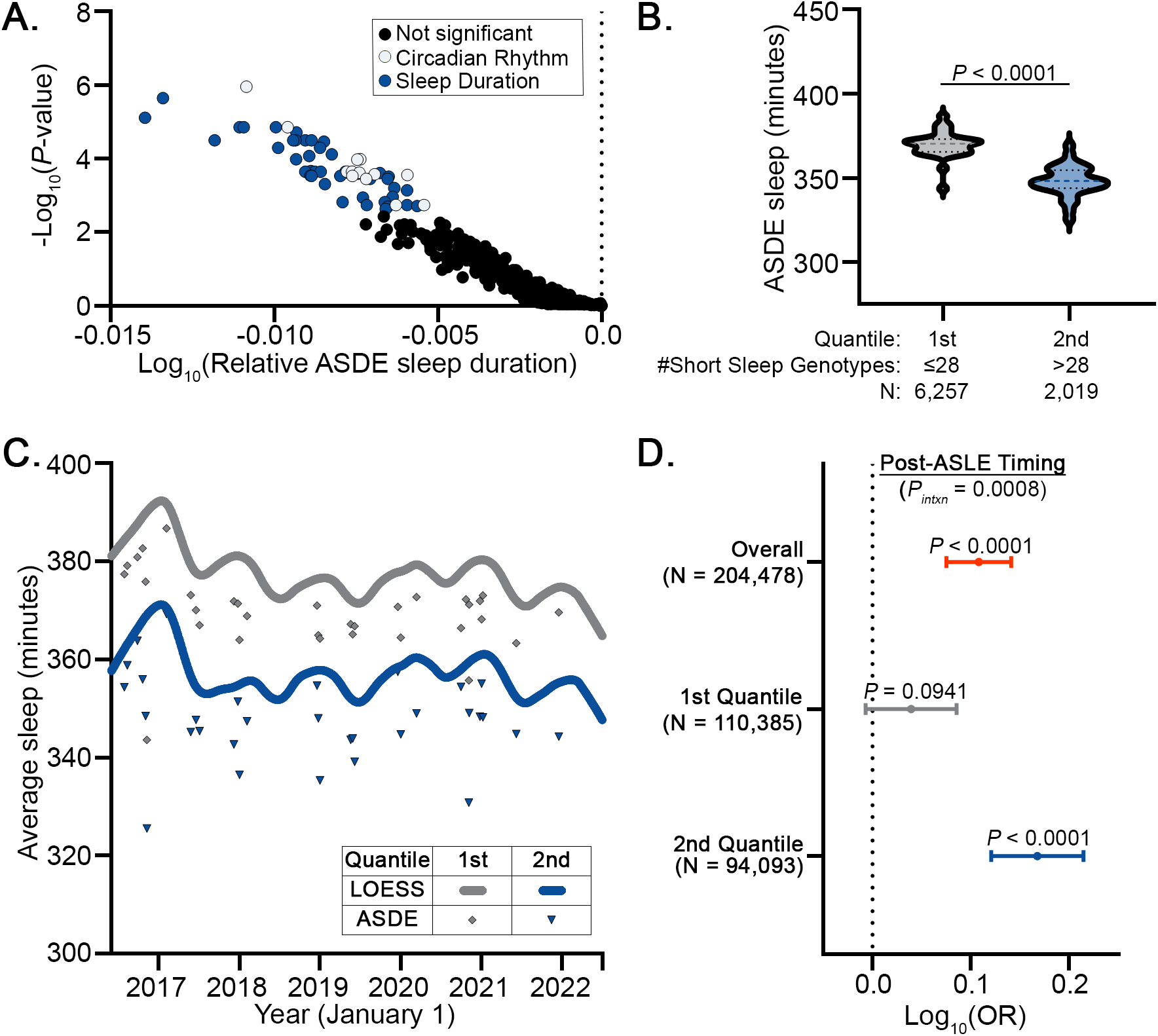
Circadian and sleep genotypes are associated with sleep duration and post-ASDE influenza risk. (**A**) Volcano plot of SNP allele combinations associated with shorter sleep duration on ASDEs. SNPs associated with significantly shorter sleep duration than the *AoU* population on ASDEs after correction for false discovery rate are highlighted. Genotyped *AoU* participants were grouped into quantiles based on their number of short sleep-associated SNP allele combinations. Sleep duration in genotyped *AoU* Fitbit population was decreased by short sleep genotype quantiles on (**B**) ASDEs and (**C**) overall. (**D**) Forest plot of Log_10_(odds ratio) of influenza diagnosis as a function of post-ASDE timing overall and grouped by short sleep genotype quantile with adjustment for age at ASDE, sex chromosome ploidy, time-weighted average weekly positive influenza tests, and genomic ancestry in a generalized estimating equation model. Comparisons in ASDE sleep duration between SNP genotypes as well as between ASDE sleep duration quantiles were performed using Mann-Whitney test. Dashed line on violin plot shows median and dotted lines show interquartile range. *P* < 0.05 was considered statistically significant. *P*_*intxn*_: interaction *P*-value (short sleep genotype quantile*post-ASDE timing).

### Synergistic interaction of ASDEs and sleep genotypes with incident influenza diagnoses

Based on our findings of the dual influence of ASDEs and sleep genotypes on sleep duration, we hypothesized that ASDEs and sleep genotypes would interact to contribute to post-ASDE incident influenza. In the subset of the genotype *AoU* population with EHR data (N = 204,478), we used generalized estimating equations (GEEs) to calculate the odds ratio of having a healthcare visit for influenza as a function of relative ASDE timing (before or after the ASDE), sleep genotype quantile, sex chromosome ploidy, genomic ancestry, and age on the date of the ASDE. To account for influenza seasonality, we also adjusted for the average weekly positive influenza tests, weighted by period length, based on public data from the U.S. Centers for Disease Control and Prevention^21^ (**Table S2**). In our model, influenza-associated healthcare visits were significantly increased after ASDEs (OR = 1.28 [1.19, 1.39], *P*-value = 1.30 x 10^-10^) in the overall population (**Table S7, Figure 3D**). In agreement with our hypothesis, the association of ASDEs with influenza visits was heightened in individuals with a greater number of short sleep genotypes (OR = 1.47 [1.32, 1.64] vs. 1.10 [0.98, 1.22], *P*-value of genotype-ASDE interaction = 0.0008, **Table S8-S10**). Together, these findings suggest that population-level acute sleep deprivation and genetic predisposition jointly influence the risk of influenza-associated healthcare visits.

## Discussion

Here, we identified the spectrum of political and non-political events associated with acute sleep deprivation in a heterogeneous sample of the American population. We demonstrated that these events, which we term ASDEs, are associated with increased incidence of influenza. We found that SNPs linked to circadian rhythm and sleep duration carry a cumulative influence on sleep duration and interact with ASDEs to jointly increase incident influenza. Taken together, this study offers needed insight into both the predisposition and health consequences of acute sleep deprivation across the American public. From a public health perspective, these findings aid in predicting future political and non-political events where sleep duration is jeopardized and where the American population is vulnerable to detrimental health outcomes.

Influenza is a recurring public health concern with between 9 and 40 million cases occurring each year in the U.S. alone^33^. Based on our findings, the upcoming elections, holidays, and major sporting events are likely to be ASDEs where an increased risk of influenza could now be predicted. Our findings should strengthen the vigilance across healthcare systems for increased influenza cases in these periods, and mitigation plans could include increased flu vaccination campaigns. Importantly, sleep deprivation has been associated with diminished anti-viral immunity following influenza vaccination^8,34^, suggesting a benefit of improved sleep hygiene proximal to vaccination, as well. These efforts may be particularly beneficial for females who, consistent with prior studies^25^, were at increased influenza risk across all time periods in our model (see **Table S10**). At the state level, we found no indication that political leanings influence the amount of sleep deprivation on ASDEs, although political leanings may vary widely across states. If future studies confirm the association of genotype with sleep duration and influenza, increased genotyping of the American public could be useful to identify those at greatest risk on future ASDEs. We do not know whether similar results would be seen in other geographic locations and countries with potentially distinct ASDEs and sociopolitical interests.

Chronic sleep deprivation has been previously associated with the risk of influenza and viral upper respiratory infections^35-39^. However, this study is the first to our knowledge identifying an association across a large and heterogeneous population between acute sleep deprivation and influenza risk. We found a robust difference in ASDE sleep duration based on an individual’s quantity of sleep and circadian-associated allelic variants, which builds upon a prior observation of the additive influence of sleep duration genotypes in a cohort with seven days of actigraphy data^40^. Our observation that genotypes associated with less habitual sleep duration are also prone to less ASDE sleep duration suggests that the relationship between chronic sleep deprivation and influenza risk may be explained by a propensity for greater acute sleep deprivation. Importantly, our model implies that the detrimental influence of sleep genotypes on influenza risk becomes robustly apparent primarily in the post-ASDE period (see **Table S10**).

Here, we focused on acute health events in the immediate aftermath of a single night of sleep deprivation. For this reason, we only examined first-time diagnoses occurring in a ten-day window following ASDEs. However, it is possible that ASDEs also pose long-term risks which could, for example, contribute to the proposed effects of major political events on mental health^41^. Longitudinal analyses of the *AoU* program will be crucial to defining these types of outcomes. Prior studies have also reported an association between major political^42^ and non-political events^43,44^ with acute cardiovascular disease. While our unbiased phenomic analysis of a smaller and younger cohort did not identify a link to cardiovascular disease, the presence of similar types of events among ASDEs raises the question of whether acute sleep deprivation may be contributing to cardiovascular disease, as well.

We acknowledge limitations to this work. This study is subject to recruitment biases of the *AoU* program, and ASDEs identified in *AoU*’s Fitbit subset may not reflect those of the larger population. In addition, while Fitbit-based sleep estimates enable population-level analyses of sleep and health^16^, they are subject to limitations and are not a substitute for gold standard polysomnography or medical-grade actigraphy^45^. Although our study demonstrates a connection between ASDEs and influenza, this finding remains associative. Our study is limited to the analysis of influenza-associated healthcare visits, which may reflect increased infection rate, greater symptom burden, or greater likelihood to seek medical attention following an ASDE. The ASDE-influenza relationship is further confounded by the fact that stress, substance use, or other unmeasured confounders may be increased around ASDEs^46,47^. Given that ASDEs coincide with major national events, it is possible that large social gatherings rather than sleep deprivation are driving the observed increases in influenza. Thanksgiving, by comparison, is associated with large gatherings in close seasonal proximity to elections but was not an ASDE; the absence of a similar increase in influenza after Thanksgiving, therefore, suggests against a major confounding contribution from large gatherings. Additionally, our incorporation of sleep genotypes strongly suggests a direct link between sleep deprivation and influenza risk, although determination of causality requires further proof. Given that sleep duration constitutes only one part of sleep health^48^, future studies should also incorporate metrics of sleep architecture beyond sleep duration.

In conclusion, this study defines the population-level landscape of ASDEs in the United States, identifies an acute association between ASDEs and influenza risk, and incorporates sleep genomics to suggest a direct link between acute sleep deprivation and influenza. We propose that these findings should guide health system preparations and public health activities around major national events.

## Data Availability

Data is available to authorized users of the All of Us Research Program Controlled Tier Dataset 7 on the Researcher Workbench at https://www.researchallofus.org/.

## Acknowledgements

We gratefully acknowledge *All of Us* participants for their contributions, without whom this research would not have been possible. We also thank the National Institutes of Health’s <u>*All of Us* Research Program</u> for making available the participant data examined in this study.

## Author Contributions

N.J.K., R.C., W.K., and S.Y.C. conceived the project and wrote the manuscript. N.J.K., W.K., N.K., J.W., P.P., I.N.C., H.R., A.S., and A.N.S. analyzed the data and prepared the figures. N.J.K., W.K., and J.W. directly accessed and verified the underlying data. S.O., C.A.M., S.M.N., and S.Y.C. supervised the project. All authors reviewed, edited, and approved the manuscript.

## Sources of Funding

This work was supported by the WoodNext Foundation (S.Y.C. and C.A.M.); NIH grants T32 HL129964 (N.J.K. and R.C.), R01 HL124021 (S.Y.C.), R01 HL122596 (S.Y.C.); R01 HL151228 (S.Y.C.); AHA grants 24SFRNCCN1276089 and 24SFRNPCN1280228 (S.Y.C.); and the McKamish Family Foundation, the Hemophilia Center of Western Pennsylvania, and the Institute for Transfusion Medicine (N.J.K. and R.C.).

## Disclosures

S.Y.C. has served as a consultant for Merck, Janssen, and United Therapeutics; S.Y.C. is a director, officer, and shareholder in Synhale Therapeutics; S.Y.C. holds research grants from United Therapeutics, Bayer, and WoodNext Foundation. S.Y.C. has filed patent applications regarding the targeting of metabolism in pulmonary hypertension. Other authors: none.

## Notes

### Author Declarations

In accordance with the All of Us Research Program, this study was exempt from human subject approval and only deidentified data were analyzed.

## References

1. Grandner MA, Fernandez F-X. The translational neuroscience of sleep: A contextual framework. Science 2021; 374(6567): 568–73.

2. Pankowska MM, Lu H, Wheaton AG, et al. Prevalence and Geographic Patterns of Self-Reported Short Sleep Duration Among US Adults, 2020. Preventing Chronic Disease 2023; 20.

3. Ford ES, Cunningham TJ, Croft JB. Trends in Self-Reported Sleep Duration among US Adults from 1985 to 2012. Sleep 2015; 38(5): 829–32.

4. Barnes CM, Drake CL. Prioritizing Sleep Health: Public Health Policy Recommendations. Perspect Psychol Sci 2015; 10(6): 733–7.

5. Spira AP, Chen-Edinboro LP, Wu MN, Yaffe K. Impact of sleep on the risk of cognitive decline and dementia. Curr Opin Psychiatry 2014; 27(6): 478–83.

6. Hertenstein E, Feige B, Gmeiner T, et al. Insomnia as a predictor of mental disorders: A systematic review and meta-analysis. Sleep Med Rev 2019; 43: 96–105.

7. Koren D, Taveras EM. Association of sleep disturbances with obesity, insulin resistance and the metabolic syndrome. Metabolism 2018; 84: 67–75.

8. Prather AA, Pressman SD, Miller GE, Cohen S. Temporal Links Between Self-Reported Sleep and Antibody Responses to the Influenza Vaccine. Int J Behav Med 2021; 28(1): 151–8.

9. Besedovsky L, Lange T, Born J. Sleep and immune function. Pflugers Arch 2012; 463(1): 121–37.

10. Lloyd-Jones DM, Allen NB, Anderson CAM, et al. Life’s Essential 8: Updating and Enhancing the American Heart Association’s Construct of Cardiovascular Health: A Presidential Advisory From the American Heart Association. Circulation 2022; 146(5).

11. Yang X, Chen H, Li S, Pan L, Jia C. Association of Sleep Duration with the Morbidity and Mortality of Coronary Artery Disease: A Meta-analysis of Prospective Studies. Heart Lung Circ 2015; 24(12): 1180–90.

12. Cappuccio FP, D’Elia L, Strazzullo P, Miller MA. Sleep duration and all-cause mortality: a systematic review and meta-analysis of prospective studies. Sleep 2010; 33(5): 585–92.

13. Banks S, Dinges DF. Behavioral and physiological consequences of sleep restriction. J Clin Sleep Med 2007; 3(5): 519–28.

14. Bjørkum AA, Griebel L, Birkeland E. Human serum proteomics reveals a molecular signature after one night of sleep deprivation. Sleep Advances 2024; 5(1).

15. Grandner MA, Lujan MR, Ghani SB. Sleep-tracking technology in scientific research: looking to the future. Sleep 2021; 44(5).

16. Zheng NS, Annis J, Master H, et al. Sleep patterns and risk of chronic disease as measured by long-term monitoring with commercial wearable devices in the All of Us Research Program. Nature Medicine 2024.

17. Metcalf M. ShowBuzzDaily. 2023. https://showbuzzdaily.com/articles/the-sked/tvratings/darilyratingsreports(accessed August 10 2024).

18. Google Trends. 2024. https://www.google.com/trends (accessed August 10 2024).

19. Bureau USC. ZIP Code Tabulation Areas. 2023. https://www.census.gov/geographies/reference-files/time-series/geo/gazetteer-files.html2024).

20. Report CP. 2022 Cook Partisan Voting Index. https://www.cookpolitical.com/cook-pvi/2022-partisan-voting-index/state-map-and-list2024).

21. CDC. Weekly U.S. influenza surveillance report. 2024. https://www.cdc.gov/flu/weekly/2024).

22. Sollis E, Mosaku A, Abid A, et al. The NHGRI-EBI GWAS Catalog: knowledgebase and deposition resource. Nucleic Acids Research 2023; 51(D1): D977–D85.

23. Bick AG, Metcalf GA, Mayo KR, et al. Genomic data in the All of Us Research Program. Nature 2024; 627(8003): 340–6.

24. Virtanen P, Gommers R, Oliphant TE, et al. SciPy 1.0: fundamental algorithms for scientific computing in Python. Nature Methods 2020; 17(3): 261–72.

25. Klein SL, Hodgson A, Robinson DP. Mechanisms of sex disparities in influenza pathogenesis. Journal of Leukocyte Biology 2012; 92(1): 67–73.

26. Black CL, O’Halloran A, Hung M-C, et al. Vital Signs: Influenza Hospitalizations and Vaccination Coverage by Race and Ethnicity—United States, 2009–10 Through 2021–22 Influenza Seasons. MMWR Morbidity and Mortality Weekly Report 2022; 71(43): 1366–73.

27. Worby CJ, Chaves SS, Wallinga J, Lipsitch M, Finelli L, Goldstein E. On the relative role of different age groups in influenza epidemics. Epidemics 2015; 13: 10–6.

28. Seabold S, Perktold J. Statsmodels: Econometric and Statistical Modeling with Python. Proceedings of the 9th Python in Science Conference; 2010. p. 92–6.

29. contributors W. Portal:Current events. 2024. https://en.wikipedia.org/wiki/Portal:Current_events (accessed September 6 2024).

30. Lipsitch M, Viboud C. Influenza seasonality: Lifting the fog. Proceedings of the National Academy of Sciences 2009; 106(10): 3645–6.

31. Ashbrook LH, Krystal AD, Fu Y-H, Ptáček LJ. Genetics of the human circadian clock and sleep homeostat. Neuropsychopharmacology 2019; 45(1): 45–54.

32. Keinan A, Clark AG. Recent Explosive Human Population Growth Has Resulted in an Excess of Rare Genetic Variants. Science 2012; 336(6082): 740–3.

33. Reed C, Chaves SS, Daily Kirley P, et al. Estimating Influenza Disease Burden from Population-Based Surveillance Data in the United States. Plos One 2015; 10(3).

34. Spiegel K, Sheridan JF, Van Cauter E. Effect of sleep deprivation on response to immunization. JAMA 2002; 288(12): 1471–2.

35. Cohen S, Doyle WJ, Alper CM, Janicki-Deverts D, Turner RB. Sleep Habits and Susceptibility to the Common Cold. Archives of Internal Medicine 2009; 169(1).

36. Jones SE, Maisha FI, Strausz S, et al. Public health impact of poor sleep on COVID-19, influenza and upper respiratory infections. Sleep Medicine 2022; 100.

37. Prather AA, Janicki-Deverts D, Hall MH, Cohen S. Behaviorally Assessed Sleep and Susceptibility to the Common Cold. Sleep 2015; 38(9): 1353–9.

38. Robinson CH, Albury C, McCartney D, et al. The relationship between duration and quality of sleep and upper respiratory tract infections: a systematic review. Fam Pract 2021; 38(6): 802–10.

39. Nieters A, Blagitko-Dorfs N, Peter HH, Weber S. Psychophysiological insomnia and respiratory tract infections: results of an infection-diary-based cohort study. Sleep 2019; 42(8).

40. Dashti HS, Jones SE, Wood AR, et al. Genome-wide association study identifies genetic loci for self-reported habitual sleep duration supported by accelerometer-derived estimates. Nat Commun 2019; 10(1): 1100.

41. Yan BW, Hsia RY, Yeung V, Sloan FA. Changes in Mental Health Following the 2016 Presidential Election. J Gen Intern Med 2021; 36(1): 170–7.

42. Mefford MT, Mittleman MA, Li BH, et al. Sociopolitical stress and acute cardiovascular disease hospitalizations around the 2016 presidential election. Proceedings of the National Academy of Sciences 2020; 117(43): 27054–8.

43. Carroll D, Ebrahim S, Tilling K, Macleod J, Smith GD. Admissions for myocardial infarction and World Cup football: database survey. BMJ 2002; 325(7378): 1439–42.

44. Phillips DP, Jarvinen JR, Abramson IS, Phillips RR. Cardiac Mortality Is Higher Around Christmas and New Year’s Than at Any Other Time. Circulation 2004; 110(25): 3781–8.

45. Haghayegh S, Khoshnevis S, Smolensky MH, Diller KR, Castriotta RJ. Accuracy of Wristband Fitbit Models in Assessing Sleep: Systematic Review and Meta-Analysis. J Med Internet Res 2019; 21(11): e16273.

46. Patrick ME, Azar B. High-Intensity Drinking. Alcohol Res 2018; 39(1): 49–55.

47. Suzuki S, Hoyt LT, Yazdani N, et al. Trajectories of sociopolitical stress during the 2020 United States presidential election season: Associations with psychological well-being, civic action, and social identities. Compr Psychoneuroendocrinol 2023; 16: 100218.

48. Buysse DJ. Sleep Health: Can We Define It? Does It Matter? Sleep 2014; 37(1): 9–17.

